# Associations between vitamin D and disease risk may be attributed to the confounding influence of adiposity during childhood and adulthood: a lifecourse Mendelian randomization study

**DOI:** 10.1101/2022.05.11.22274956

**Authors:** Tom G Richardson, Grace M Power, George Davey Smith

## Abstract

**Background:** Vitamin D supplements are widely prescribed to help reduce disease risk. However, this strategy is based on findings using conventional epidemiological methods which are prone to confounding and reverse causation.

**Methods:** In this short report, we leveraged genetic variants which differentially influence body size during childhood and adulthood within a multivariable Mendelian randomization (MR) framework, allowing us to separate the genetically predicted effects of adiposity at these two timepoints in the lifecourse.

**Results:** Using data from the Avon Longitudinal Study of Parents and Children (ALSPAC), there was strong evidence that higher childhood body size has a direct effect on lower vitamin D levels in early life (mean age: 9.9 years, range=8.9 to 11.5 years) after accounting for the effect of the adult body size genetic score (Beta=-0.32, 95% CI=-0.54 to -0.10, P=0.004). Conversely, we found evidence that the effect of childhood body size on vitamin D levels in midlife (mean age: 56.5 years, range=40 to 69 years) is putatively mediated along the causal pathway involving adulthood adiposity (Beta=-0.17, 95% CI=-0.21 to -0.13, P=4.6×10^−17^).

**Conclusions:** Our findings have important clinical implications in terms of the causal influence of vitamin D deficiency on disease risk. Furthermore, they serve as a compelling proof of concept that the timepoints across the lifecourse at which exposures and outcomes are measured can meaningfully impact overall conclusions drawn by MR studies.

## Introduction

Associations between vitamin D deficiency and disease risk have been widely reported by conventional epidemiological studies, including diseases which typically have a late-onset over the lifecourse, such as coronary artery disease, but also those which may be diagnosed in early life such as type 1 diabetes (T1D). As a result, vitamin D supplements are widely prescribed with an estimated 18% of adults in the USA reportedly taking supplements daily (Rooney *et al*., 2017). However, there is increasing evidence from the literature suggesting that vitamin D supplements may be ineffective at reducing disease risk in the population (Murai *et al*., 2021), and although there are notable exceptions (e.g. multiple sclerosis (Mokry *et al*., 2015, Vandebergh *et al*., 2022)), this raises uncertainty into the causal effects of vitamin D levels on many disease outcomes. Furthermore, these conventional associational studies may have been susceptible to bias, given that vitamin D levels are known to differ amongst individuals based on various lifestyle factors, including their body mass index (BMI), as well as being prone to reverse causation, for example due to inflammatory processes which are known to lower vitamin D levels (Preiss and Sattar, 2019).

An approach to mitigate the influence of these sources of bias is Mendelian randomization (MR), a causal inference technique which exploits the random allocation of genetic variants at birth to evaluate the genetically predicted effects of modifiable exposures on disease outcomes and circulating biomarkers (Davey Smith and Ebrahim, 2003, Richmond and Davey Smith, 2022). For example, MR has been applied in recent years to suggest that vitamin D supplements are unlikely to have a beneficial effect on risk of type 1 diabetes (Manousaki *et al*., 2021). We previously extended the application of MR to investigate epidemiological hypotheses in lifecourse setting (known to as ‘lifecourse MR’), by deriving sets of genetic variants to separate the independent effects of body size during childhood and adulthood within a multivariable framework (Richardson *et al*., 2020). Applying this approach has highlighted the putative causal role that early life adiposity may have on outcomes such as T1D risk (Richardson *et al*., 2022) and cardiac structure (O’Nunain et al (in press)). In contrast, we have demonstrated that it’s influence on other disease outcomes (e.g. cardiovascular disease (Power *et al*., 2021)) is likely attributed to the long-term consequence of remaining overweight into later life. Whilst these applications serve as powerful examples of lifecourse MR as an approach to separate the effects of the same exposure based on data derived from early and later life, it has yet to be applied to the same outcome when measured at different timepoints in the lifecourse.

## Results

In this study, we applied lifecourse MR to investigate the independent effects of childhood and adult body size has a largely indirect influence on 25-hydroxyvitamin D (25OHD) levels measured during childhood (mean age: 9.9 years, range=8.9 to 11.5 years) using individual-level data from the Avon Longitudinal Study of Parents and Children (ALSPAC) (Boyd *et al*., 2013, Fraser *et al*., 2013) and during adulthood (mean age: 56.5 years, range=40 to 69 years) using summary-level data based on individuals from the UK Biobank (UKB) study (Manousaki *et al*., 2020). Firstly, using data derived from the ALSPAC study, our analyses indicated that childhood body size directly influences vitamin D levels during childhood after accounting for the adult body size genetic instrument in our model (Beta=-0.32 standard deviation (SD) change in natural log-transformed 25OHD per change in body size category, 95% CI=-0.54 to -0.10, P=0.004) (**Figure 1A & Figure 1B**). Using data from the UKB study, evidence of an effect of higher childhood body size on adulthood measured 25OHD levels (Beta=-0.14, 95% CI=-0.10 to -0.03, P=2.4×10^−4^) attenuated in a multivariate setting upon accounting for adulthood body size (Beta=0.04, 95% CI=-0.01 to 0.08, P=0.10). In contrast, strong evidence of an effect for higher adult body size on lower 25OHD levels measured in adulthood was found in the multivariable model (Beta=-0.17, 95% CI=-0.21 to -0.13, P=4.6×10^−17^) (**Figure 1C**). This suggests that childhood body size has an indirect influence of 25OHD levels in adulthood, likely due to its persistent effect throughout the lifecourse (**Figure 1D**).

**Figure 1.**
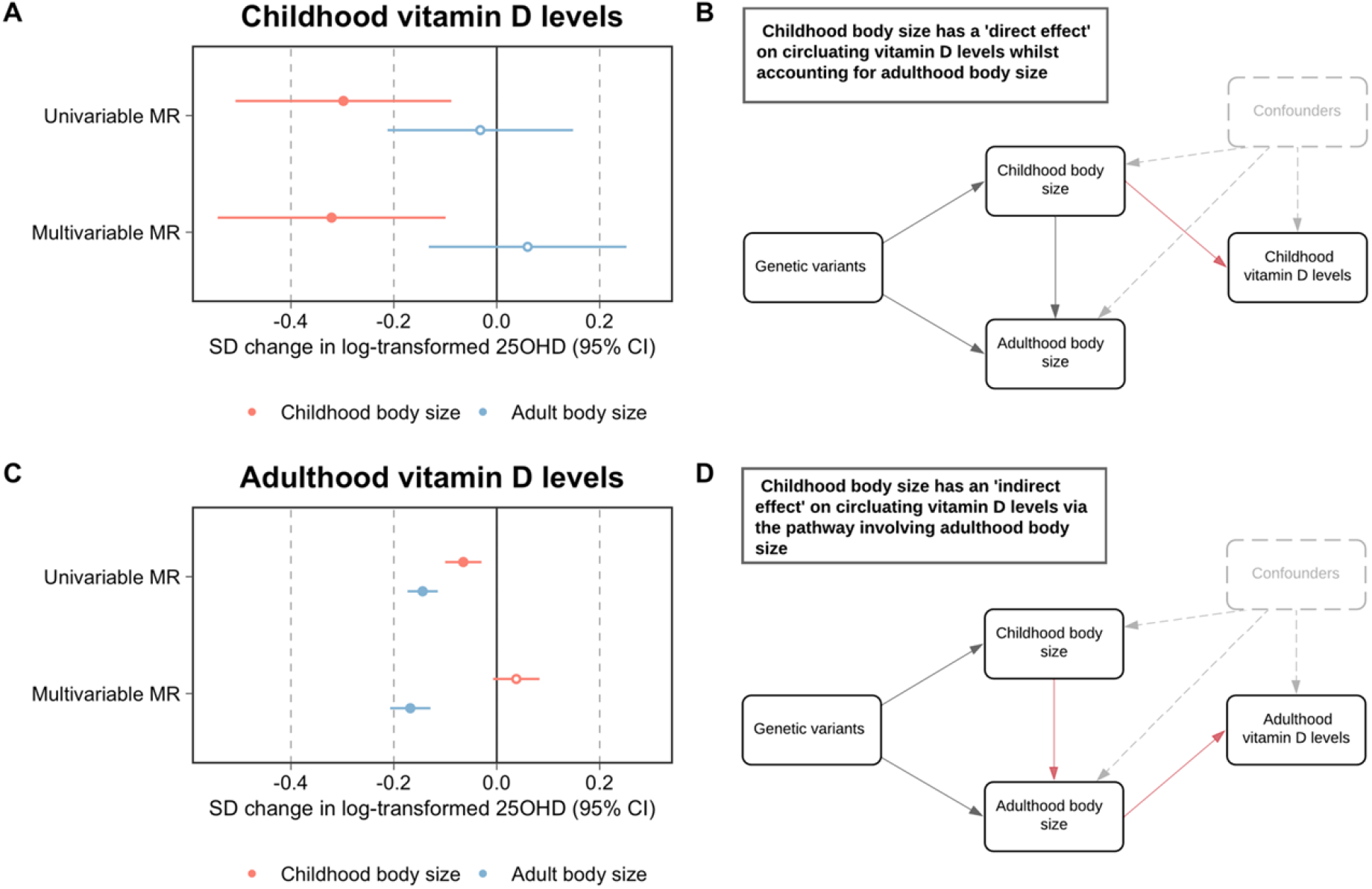
A) A forest plot illustrating the direct effect of childhood body size on circulating vitamin D levels measured during childhood (mean age: 9.9 years) using participant data from the Avon Longitudinal Study of Parents and Children (ALSPAC) with the corresponding schematic diagram for this finding being located in panel B). C) A forest plot depicting the indirect effect of childhood body size on adulthood measured vitamin D levels using data from the UK Biobank study (mean age: X years) as described in the schematic diagram presented in panel D). MR – Mendelian randomization.

## Discussion

Our findings suggest that increased adiposity exerts a strong effect on lower vitamin D levels during both childhood and adulthood. Separating causal from confounding factors, particularly in a lifecourse setting, would have been extremely challenging to disentangle without the use of genetic variants as achieved in this study. In contrast, appropriately accounting for confounding factors in an conventional epidemiological setting is notoriously challenging, with previous studies reporting evidence of association between vitamin D and T1D in early life even after adjusting for factors such as birthweight (Hypponen *et al*., 2001). Taken together with evidence from previous MR studies, which have found that childhood body size (Richardson *et al*., 2022), but not vitamin D levels (Manousaki *et al*., 2021), increases risk of T1D, our results suggest that adiposity may have acted as a confounding factor on the observed association between vitamin D and T1D. These findings therefore have important clinical implications in terms of whether patients newly diagnosed with T1D should be prescribed vitamin D supplements or not.

Effect estimates derived from MR studies are conventionally interpreted as ‘lifelong effects’ given that the germline genetic variants harnessed by this approach as instrumental variables are typically fixed at conception. The results of this study serve as a compelling demonstration that the timepoint at which both exposures and outcomes are measured across the lifecourse can meaningfully impact conclusions drawn by MR investigations. Whilst the childhood and adult body size genetic scores were used in this study as a proof of concept, our findings also help to further validate their utility in separating the temporal effects of adiposity within a lifecourse MR framework. Overall, these findings emphasise the importance of future MR studies taking further consideration into the age of participants that their genetic estimates are based on, as well as the age at which cases are diagnosed for disease outcome estimates, before interpreting and drawing conclusions from their findings. Furthermore, our results highlight the importance of conducting genome-wide association studies on populations of different age groups to help uncover time-varying genetic effects scattered throughout the human genome. Findings from these endeavours should facilitate insight into the direct and indirect effects of modifiable early life exposures using approaches such as lifecourse MR. This may help to elucidate the critical timepoints whereby conferred risk by these exposures on disease outcomes starts to become immutable, which has important implications for improving patient care in a clinical setting.

## Materials and Methods

ALSPAC is a population-based cohort investigating genetic and environmental factors that affect the health and development of children. The study methods are described in detail elsewhere (Boyd *et al*., 2013, Fraser *et al*., 2013). In brief, 14,541 pregnant women residents in the former region of Avon, UK, with an expected delivery date between April 1, 1991 and December 31, 1992, were eligible to take part in ALSPAC. Detailed phenotypic information, biological samples and genetic data which have been collected from the ALSPAC participants are available through a searchable data dictionary (http://www.bris.ac.uk/alspac/researchers/our-data/). Written informed consent was obtained for all study participants. Ethical approval for this study was obtained from the ALSPAC Ethics and Law Committee and the Local Research Ethics Committees. Measures of 25-hydroxyvitamin D (25OHD) levels were obtained from non-fasting blood samples taken from ALSPAC participants at mean age 9.9 years (range=8.9 to 11.5 years) which were log transformed to ensure normality.

Derivation of genetic instruments for childhood and adulthood body size have been described in detail previously (Richardson *et al*., 2020). In brief, genome-wide association studies (GWAS) were conducted on 463,005 UK Biobank (UKB) participants (mean age: 56.5 years, range=40 to 69 years) who had both reported their body size at age 10 as well as had their BMI clinically measured. Genetic instruments were identified from these analyses (based on P<5×10^−8^) and the resulting genetic score for childhood body size has been validated using measured childhood BMI in ALSPAC (Richardson *et al*., 2020), the Young Finns Study (Richardson *et al*., 2021) and the Trøndelag Health (HUNT) study (Brandkvist *et al*., 2020). Genetic estimates on adulthood 25OHD were obtained from a previously conducted GWAS in UKB (Manousaki *et al*., 2020). Despite having overlapping samples when analysing our body size instruments against the adulthood measure of vitamin D, there was little evidence of inflated type 1 error rates (based on the calculator at https://sb452.shinyapps.io/overlap) (Burgess *et al*., 2016).

Mendelian randomization (MR) analyses to estimate genetically predicted effects on childhood 25OHD were conducted in a one-sample setting using individual level data from ALSPAC after generating genetic risk scores for our body size instruments with adjustment for age and sex. MR analyses to estimate effects on adulthood 25OHD were undertaken in a two-sample setting using the inverse variance weighted (IVW) method (Burgess *et al*., 2013), as this allowed us to additionally perform analyses using the weighted median and MR-Egger methods (Bowden *et al*., 2015, Bowden *et al*., 2016). (**Supplementary Table 1**). Multivariable MR were performed in one- and two-sample settings respectively for childhood and adulthood measures of 25OHD (Sanderson *et al*., 2019) (**Supplementary Table 2**).

## Data Availability

All individual level data analysed in this study can be accessed via an approved application to ALSPAC (http://www.bristol.ac.uk/alspac/researchers/access/). Summary-level data on adulthood vitamin D levels can be accessed publicly via the OpenGWAS (https://gwas.mrcieu.ac.uk/).

## Consent

Written informed consent was obtained for all study participants. Ethical approval for this study was obtained from the ALSPAC Ethics and Law Committee and the Local Research Ethics Committees.

## Acknowledgements

We are extremely grateful to all the families who took part in this study, the midwives for their help in recruiting them and the whole ALSPAC team, which includes interviewers, computer and laboratory technicians, clerical workers, research scientists, volunteers, managers, receptionists and nurses. The UK Medical Research Council and Wellcome (Grant ref: 217065/Z/19/Z) and the University of Bristol provide core support for ALSPAC. Consent for biological samples has been collected in accordance with the Human Tissue Act (2004). GWAS data was generated by Sample Logistics and Genotyping Facilities at Wellcome Sanger Institute and LabCorp (Laboratory Corporation of America) using support from 23andMe.

This research was conducted at the NIHR Biomedical Research Centre at the University Hospitals Bristol NHS Foundation Trust and the University of Bristol. The views expressed in this publication are those of the author(s) and not necessarily those of the NHS, the National Institute for Health Research or the Department of Health. This publication is the work of the authors and TGR will serve as guarantor for the contents of this paper.

